# Postoperative radiotherapy timing, molecular subgroups and treatment outcomes of Thai pediatric patients with medulloblastoma

**DOI:** 10.1101/2022.07.08.22277408

**Authors:** Thitiporn Jaruthien, Chonnipa Nantavithya, Sakun Santisukwongchote, Shanop Shuangshoti, Piti Techavichit, Darintr Sosothikul, Jiraporn Amornfa, Kanjana Shotelersuk

## Abstract

**Introduction:** Medulloblastoma (MB) is the most common childhood malignant brain tumor worldwide. Recently, molecular classification was established and started to play a role in the management of MB; however, studies involving molecular defined MB in Southeast Asia have been limited. We aimed to describe, and correlate clinical characteristics and molecular subgroups with therapeutic outcomes of Thai pediatric patients with MB.

**Materials and Methods:** Pediatric MB patients treated at King Chulalongkorn Memorial Hospital in Thailand from 2008 to 2018 were recruited. Patients were classified by clinical characteristics into standard- and high-risk groups, which determined treatment regimen. Retrospectively, available tumor tissues were classified into 3 molecular subgroups using immunohistochemistry: 1) WNT, 2) SHH, and 3) non-WNT/non-SHH. The primary outcome was 5-year overall survival (OS). Risk factors associated with OS were analyzed using cox regression analysis.

**Results:** Fifty-three Thai pediatric patients with MB were enrolled. The median follow-up time was 60 months. The 5-year OS for all patients, and patients with standard-risk and high-risk were 74.2%, 76.3% and 71.4%, respectively. Tumor tissues of 24 patients were available, of which 23 could be molecularly classified. Two, one and 20 were in the WNT, SHH and non-WNT/non-SHH subtypes with 5-year OS of 100%, 100% and 78.9%, respectively. Using multivariate analysis, the interval of more than 8 weeks between surgery and radiotherapy was significantly correlated with a decrease in the 5-year OS.

**Conclusion:** Interval between surgery and radiotherapy within 8 weeks was associated with good therapeutic outcomes among Thai pediatric patients with MB. Simplified molecular subtyping combined with clinical characteristics is practical in risk classification of patients with MB in institutes with limited resources.

## Introduction

Cancer is the leading cause of death in children worldwide. The most common cancers in children are leukemia, followed by central nervous system (CNS) tumors [1]. Medulloblastoma (MB) is the most common childhood malignant brain tumor, accounting for 20% of all pediatric brain malignancies [2].

Patients with MB are conventionally divided into two clinical risk groups using clinical information such as age, postoperative residual tumor, metastatic disease, and histopathology: 1) standard-risk, and 2) high-risk groups. The current standard of treatment consists of maximal safe resection followed by craniospinal irradiation and chemotherapy adjusted according to the risk groups. Recently, surgical procedures, diagnostic imaging, radiotherapy techniques and the implementation of systemic therapy have improved compared to the past, thus, the outcome of treatment for MB patients has also improved. Current studies have reported 5-year OS for standard-risk group and high-risk group to be 70-85% and 60-70%, respectively [3-5].

Several studies had identified prognostic factors affecting outcome of MB patients. These included not only tumor-intrinsic factors such as histology and molecular features but also demographic data such as age and sex, and treatment-related features such as extent of surgery, and time interval between surgery and radiotherapy [6, 7]. The benefit of knowing prognostic factors is a key in optimizing treatment to improve the treatment outcome as well as reduce the long-term toxicities in selected patients.

At present, we have better understanding of molecular and genetic behaviors of this tumor. The concept for CNS tumor diagnoses has been shifted by using molecular parameters in addition to histology to define the tumor entities. According to the most recent 2021 World Health Organization (WHO) Classification of Tumors of the Central Nervous System [8], four main molecular subgroups of MB patients are classified: 1) WNT, 2) SHH with TP53-wildtype, 3) SHH with TP53-mutant, and 4) non-WNT/non-SHH. These four distinct subgroups have different prognosis and outcomes in various ethnicities. WNT patients are considered to have the best prognosis. Thus, the current consensus has been used in many recent research studies to guide treatment in the future. For example, two open studies, PNET5 (NCT02066220) and SJMB12 (NCT01878617), are evaluating a reduction of craniospinal irradiation dose in MB patients based on both the clinical risk and molecular subtypes.

Although molecular subgroups are proposed as an important prognostic factor which can potentially change the treatment paradigm in the near future, unfortunately there are limited number of MB studies in Southeast Asia [9, 10]. To date, there is only one Thai study that assessed the clinical outcome of MB treatment among pediatric patients based on their molecular subgroups [11]. Limited data regarding the prognosis of molecular defined MB in Thai patients because molecular genetic testing is not available at all centers. Therefore, we aimed to conduct a retrospective study to identify the prognostic factors, including molecular pathology, of the treatment outcome of MB in Thai pediatric patients.

## Materials and methods

### Study population

Consecutive patients aged under 16 years who were diagnosed with MB by histopathology at the King Chulalongkorn Memorial Hospital from 2008 to 2018 were retrospectively reviewed. All underwent surgical tumor removal followed by craniospinal irradiation (CSI). Patients with incomplete treatment or missing medical records were excluded from the study. Histopathology was stratified into classic, desmoplastic/nodular, extensively nodular, and large cell/anaplastic. The high-risk group was defined as the age at diagnosis of less than 3 years, have residual disease after resection larger than 1.5 cm^2^, have evidence of metastases at diagnosis or histopathology of large cell/anaplastic type. Patients without any of these conditions were categorized as standard-risk group. The study protocol was approved by the Research Ethics Committee of King Chulalongkorn Memorial Hospital and written informed consent was waived due to retrospective study.

### Treatment and Follow-up

All patients received a pretreatment evaluation including general physical examination, imaging of the brain and the whole spine, and lumbar puncture to study cerebrospinal fluid cytology. Patients were treated according to the Thai Pediatric Oncology Group (ThaiPOG) protocol [12]. Briefly, all patients underwent maximal safe resection of the primary tumor followed by CSI and involved field and/or posterior fossa boost and chemotherapy. For the standard risk group, the CSI dose was 23.4 Gy followed by either whole posterior fossa irradiation to at least 36 Gy and a tumor bed boost or irradiation of the tumor bed alone to 54-55.8 Gy. For the high-risk group, the CSI dose was 36 Gy followed by either whole posterior fossa boost or a tumor bed boost up to 54-55.8 Gy. Chemotherapy was administered concurrently with vincristine 1.5 mg/m^2^ weekly during radiotherapy (RT) for six doses and adjuvantly with cyclophosphamide plus vincristine alternate with carboplatin plus etoposide for a total of ten cycles unless the condition of the patients did not allow chemotherapy to be given. Following treatment completion, patients were followed with physical examination and serial MRI of the brain and the whole spine every 3 months for the first 2 years, every 6 months for the 3^rd^ and 4^th^ years, and yearly thereafter. Acute toxicities were graded according to the Common Terminology Criteria for Adverse Events (CTCAE) version 3.0.

### Molecular subgroup classification

MB tumor specimens from the Department of Pathology were reviewed, and the molecular subtyped determined by the method previously described [13]. In brief, a panel of 5 immunostains (Beta-catenin, GAB, YAP, NGFR, OTX2) was first applied. Cases with the classic immunoprofile were categorized into, 1) WNT, 2) SHH, and 3) non-WNT/non-SHH (group 3 and group 4). CTNNB1 mutation test was additionally performed in tumors with negative nuclear Beta-catenin but the other immunohistochemical markers perfectly matched with WNT MB. Cases were considered unclassified molecular subtype when the 5 immunostains did not fit with any of the molecular subtypes, including the WNT subtype with negative nuclear Beta-catenin.

### Statistical analysis

Overall survival (OS) was defined as the time from diagnosis until death by any cause. Event free survival (EFS) was defined as the time from diagnosis until disease progression, disease recurrence or death. The date of tumor resection was determined as the date of diagnosis. Median was compared using Wilcoxon rank sum test. Comparing proportion using Chi-square or fisher exact test. OS and EFS were assessed using the Kaplan-Meier method. Log rank test was used to compare survival between the 2 groups. Risk factors associated with OS and EFS were assessed using Cox regression. Covariates with p<0.2 in the univariate models were analyzed in the multivariate models. Statistical significance was defined as p-values <0.05. Statistical analysis was performed using STATA software, version 15.1 (Stata Corp., College Station, Texas).

## Results

### Patient and Tumor Characteristics

From 2008 to 2018, 56 patients were eligible but three patients were excluded due to either incomplete treatment or missing treatment information, resulting in a remaining total of 53 patients. The median follow-up time was 60 months, with a range of 8 months to 13 years. Median age of all patients at the time of diagnosis was 7 years (interquartile range 5-10 years). The most common histology was the classic type. Three patients did not receive chemotherapy due to poor performance status and refused to be treated with chemotherapy, while 50 patients received a combination of craniospinal irradiation and chemotherapy. Six patients received neoadjuvant chemotherapy. Gross total resection (GTR) or near total resection (NTR) was performed on 88.7% of the patients. The demographic, tumor and treatment characteristics of the patients are summarized in Table 1.

**Table 1.** Demographic and clinical characteristics of the 53 pediatric patients with medulloblastoma.

|  | **Total**<br>**(N=53)** | **Alive**<br>**(N=37)** | **Dead**<br>**(N=16)** | **P-value** |
| --- | --- | --- | --- | --- |
| **Median (IQR) age** | 7 (5-10) | 8 (4-10) | 7 (6-12.5) | 0.73 |
| **Male** | 31 (58.5) | 19 (51.4) | 12 (75) | 0.11 |
| **Location** |  |  |  | 0.52 |
| **Vermis** | 51 (96.2) | 36 (97.3) | 15 (93.8) |  |
| **Hemisphere** | 2 (3.8) | 1 (2.7) | 1 (6.2) |  |
| **Histology** |  |  |  | 0.94 |
| **Classic** | 38 (71.7) | 27 (73) | 11 (68.7) |  |
| **Desmoplastic nodular** | 4 (7.6) | 3 (8.1) | 1 (6.3) |  |
| **Large/anaplastic** | 2 (3.8) | 1 (2.7) | 1 (6.3) |  |
| **Unknown** | 9 (16.9) | 6 (16.2) | 3 (18.7) |  |
| **Clinical Risk** |  |  |  | 0.93 |
| **Standard** | 26 (49.1) | 18 (48.7) | 8 (50) |  |
| **High** | 27 (50.9) | 19 (51.3) | 8 (50) |  |
| **High risk criteria** |  |  |  | 0.75 |
| **< 3 years** | 2 (7.4) | 1 (5.3) | 1 (12.5) |  |
| **M1** | 17 (63) | 12 (63.2) | 5 (62.5) |  |
| **>1.5 cm<sup>2</sup>** | 1 (3.7) | 1 (5.3) | 0 |  |
| **Age+M1** | 2 (7.4) | 2 (10.5) | 0 |  |
| **M1+residual>1.5 cm<sup>2</sup>** | 5 (18.5) | 3 (15.8) | 2 (25) |  |
| **Treatment** |  |  |  | 0.29 |
| **Radiotherapy with chemotherapy** | 50 (94.3) | 36 (97.3) | 14 (87.5) |  |
| **Radiotherapy alone** | 3 (5.7) | 1(2.7) | 2 (12.5) |  |
| **Neoadjuvant chemotherapy** | 6 (11.3) | 4 (10.8) | 2 (12.5) | 0.86 |
| **Extent of tumor removal** |  |  |  | 0.75 |
| **GTR/NTR** | 47 (88.7) | 33 (89.2) | 14 (87.5) |  |
| **STR** | 5 (9.4) | 3 (8.1) | 2 (12.5) |  |
| **Partial tumor removal** | 1 (1.9) | 1 (2.7) | 0 |  |

Abbreviation: IQR; interquartile range, GTR; gross total resection, NTR; near total resection STR; subtotal resection

## Outcomes

Of the 16 patients (30.2%) who died, eight (50%) were from the standard-risk group. The 5-year OS for all patients was 74.2%. The 5-year OS were 76.3% and 71.4% in patients from the standard-risk and high-risk groups, respectively. There was no statistically significant difference in OS between the 2 groups (P = 0.79). Nine patients had recurrence before death. The 5-year EFS were 73.1% and 66% in patients from the standard-risk and high-risk groups, respectively. The OS and EFS are shown in Fig 1. Concerning RT doses, all patients received CSI dose as planned. For tumor bed boost, whole posterior fossa boost to 54 Gy was given in 2/26 (7.7%) and 14/27 (51.9%) in the standard-risk and high-risk groups, respectively. Local recurrence was documented in 12 cases. Six patients who had local recurrence were in the high-risk group, three receiving posterior fossa boost while the other three receiving only tumor bed boost to 54 Gy.

**Fig 1.**
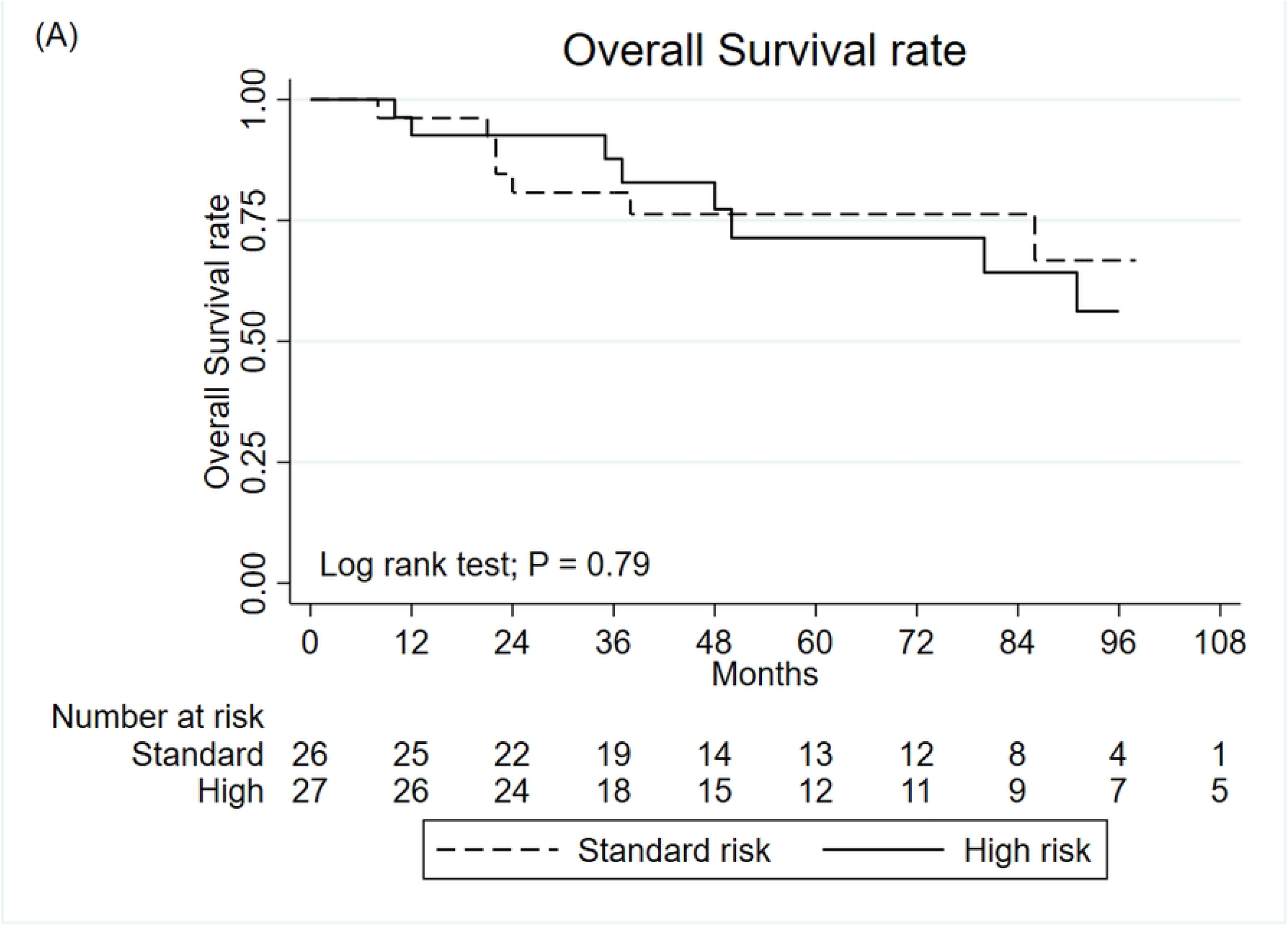
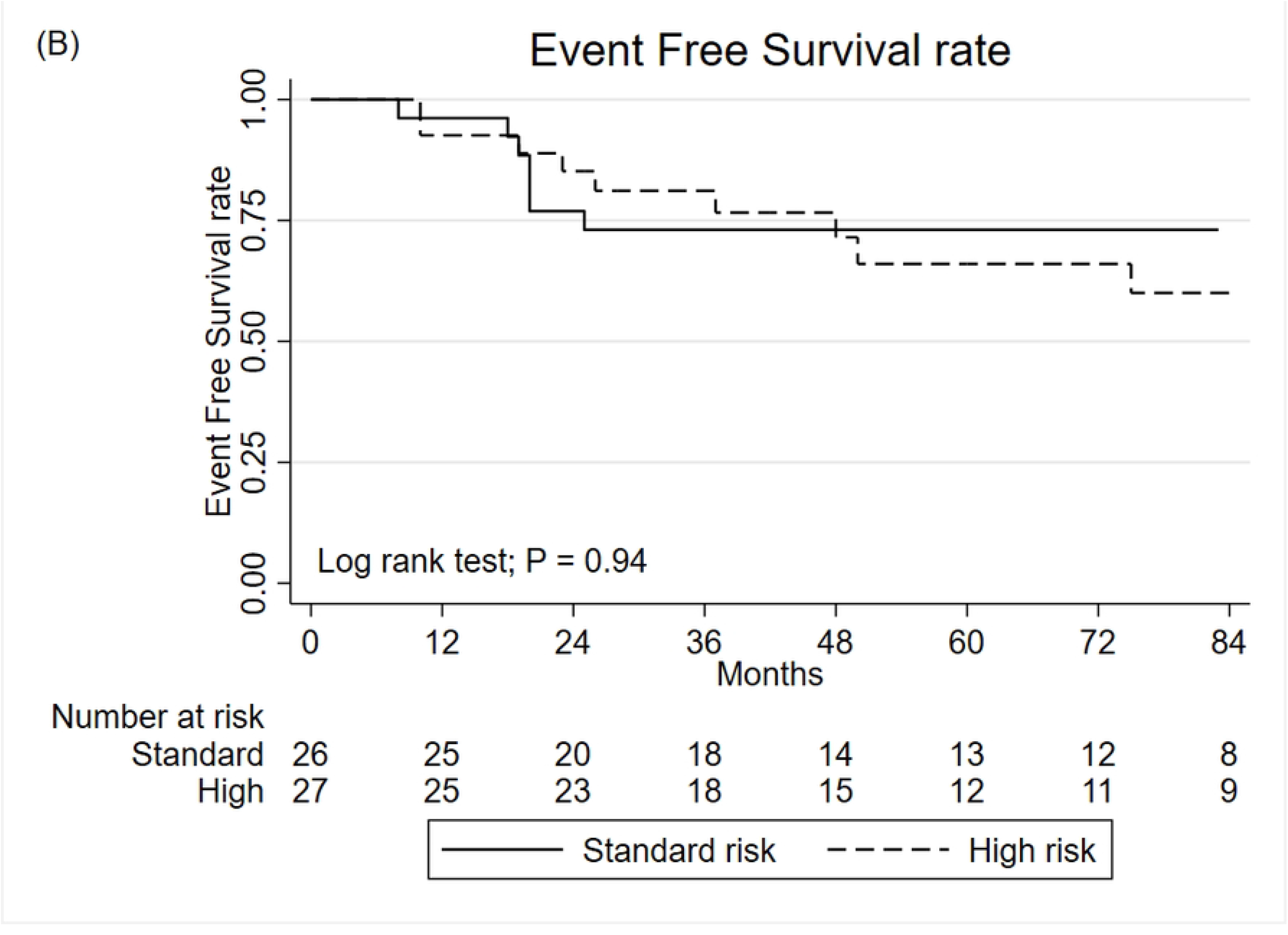
The Kaplan-Meier curve for overall survival (A) and event free survival (B) of pediatric patients from the standard risk and high risk medulloblastoma groups.

### Prognostic factors affecting overall survival rates

In the univariate analysis, sex, tumor location, extent of resection and histological subtypes did not affect OS. In the multivariate analysis, the interval of more than 8 weeks between surgery and RT was significantly associated with a decrease in OS. The results of the univariate and multivariate analyses are summarized in Table 2. The tumor tissues of 24 cases could be retrieved and were reviewed by pathologists for molecular subtype classification. The rest of the cases did not yield adequate material for analysis. One in 24 patients was considered unclassified subtype by immunohistochemistry. Demographic data, molecular and histologic subtypes of 23 MB patients are shown in S1 Table. The predominant subtype (20/23; 86.9%) was non-WNT/non-SHH. Two cases were WNT subtype, 1 case was SHH subtype. The correlation between molecular subtype, clinical risk and histopathology are summarized in Table 3.

**Table 2.** The association analysis of various factors and the overall survival of the medulloblastoma pediatric patients.

|  | N | Univariate |  | Multivariate |  |
| --- | --- | --- | --- | --- | --- |
|  |  | HR (95%CI) | P-value | HR (95%CI) | P-value |
| **Sex** |  |  |  |  |  |
| **Female** | 22 | Ref |  |  |  |
| **Male** | 31 | 2.01 (0.64-6.28) | 0.23 |  |  |
| **Location** |  |  |  |  |  |
| **Vermis** | 51 | Ref |  |  |  |
| **Hemisphere** | 2 | 4.17 (0.52-33.11) | 0.17 | 4.82 (0.58-39.78) | 0.14 |
| **Extent of resection** |  |  |  |  |  |
| **GTR/NTR** | 47 | Ref |  |  |  |
| **STR** | 5 | 1.81 (0.4-8.21) | 0.44 |  |  |
| **Partial tumor removal** | 1 | N/A |  |  |  |
| **Histologic type** |  |  |  |  |  |
| **Classic** | 38 | Ref |  |  |  |
| **Desmoplastic/nodular** | 4 | 0.57 (0.07-4.52) | 0.6 |  |  |
| **Large/anaplastic** | 2 | 1.63 (0.21- 12.83) | 0.64 |  |  |
| **Interval between surgery and RT** |  |  |  |  |  |
| **< 4 weeks** | 8 | Ref |  |  |  |
| **≥ 4 weeks** | 45 | 0.72 (0.2-2.56) | 0.61 |  |  |
| **Interval between surgery and RT** |  |  |  |  |  |
| **< 8 weeks** | 40 | Ref |  |  |  |
| **≥ 8 weeks** | 13 | 3.15 (1.16-8.52) | 0.02 | 3.25 (1.19-8.85) | 0.02 |

Abbreviation: GTR; gross total resection, NTR; near total resection, STR; subtotal resection, RT; radiotherapy

**Table 3.** Molecular classification of the medulloblastoma pediatric patients.

| Patient characteristics | Molecular subgroups |  |  | Total (n=23) |
| --- | --- | --- | --- | --- |
|  | WNT | SHH | nonWNT/nonSHH |  |
| Clinical risk |  |  |  |  |
| Standard risk | 2 | 0 | 9 | 11 |
| High risk | 0 | 1 | 11 | 12 |
| Histopathology |  |  |  |  |
| Classic | 2 | 0 | 20 | 22 |
| Desmoplastic/ nodular | 0 | 1 | 0 | 1 |

Four patients from the non-WNT/non-SHH group died. All patients from WNT and SHH subtype were completed the standard treatment as planned and remained disease free until the last follow-up visit. The 5-year OS were 100%, 100% and 78.9% for WNT, SHH and non-WNT/non-SHH subtypes, respectively. The 5-year EFS were 100%, 100% and 74.3% for WNT, SHH and non-WNT/non-SHH subtypes, respectively (Fig 2).

**Fig 2.**
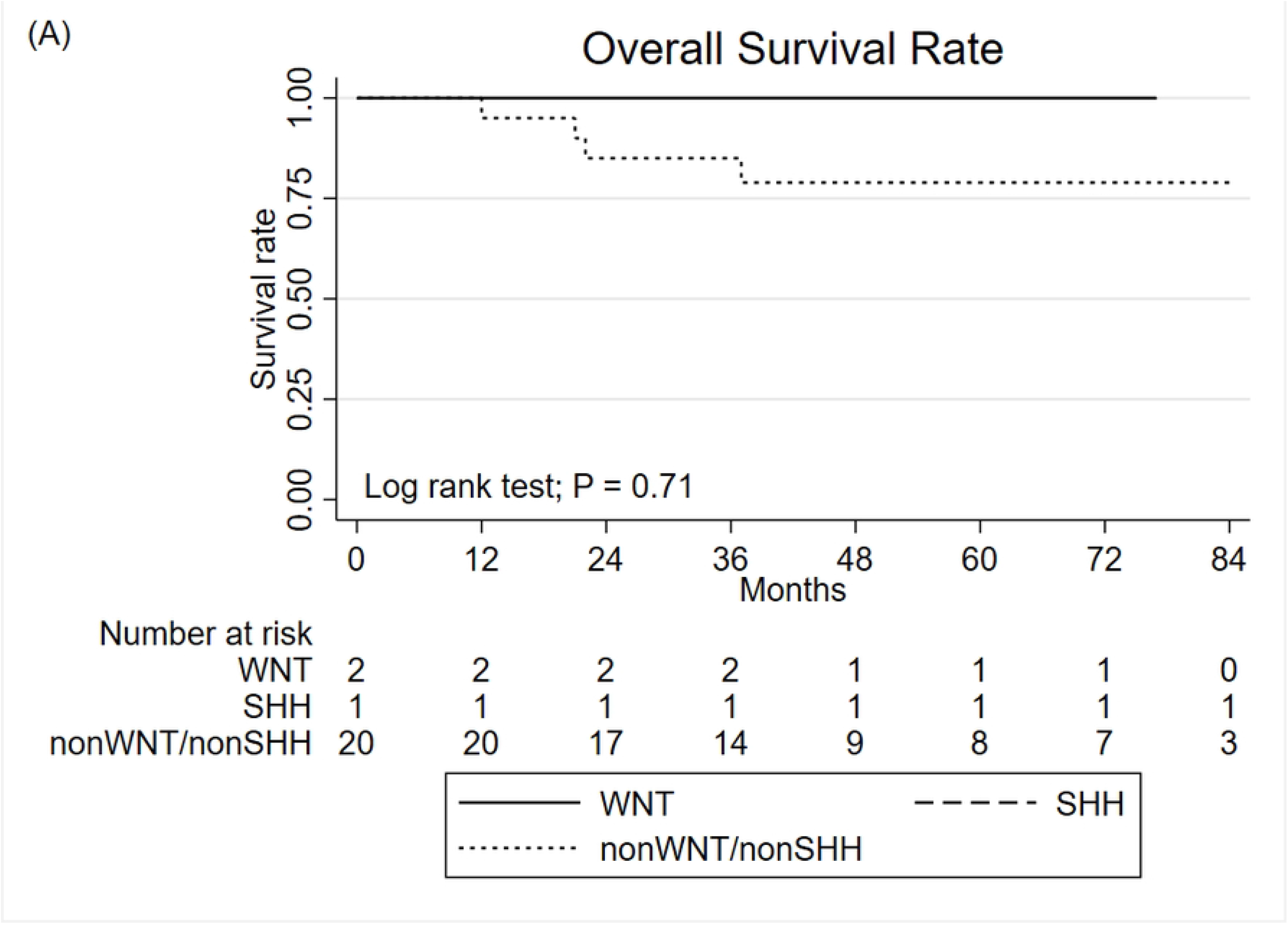
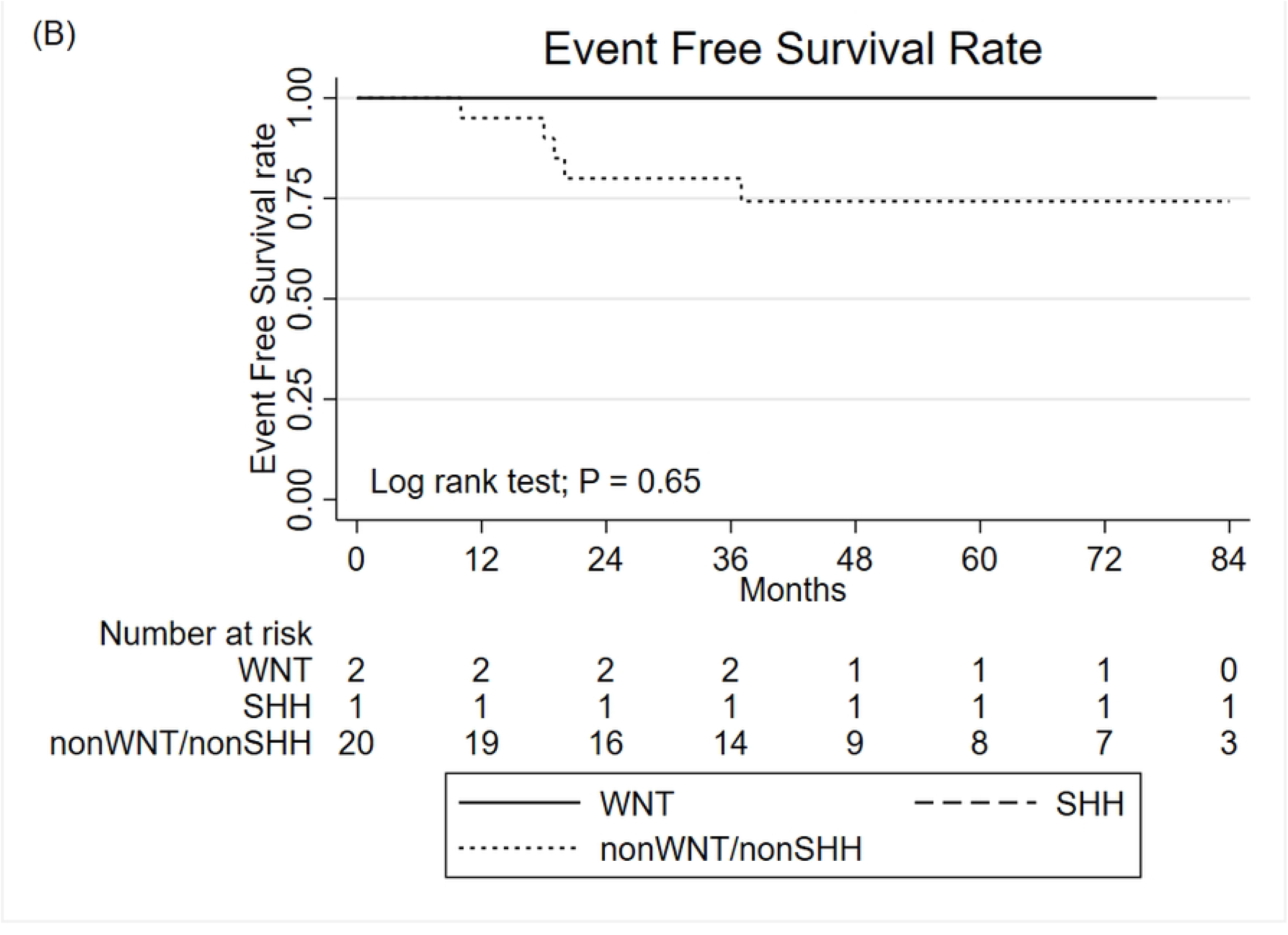
The Kaplan-Meier curve for overall survival (A) and event free survival (B) of medulloblastoma by molecular subtypes.

### Toxicities

Acute hemato-toxicities were recorded. Grade 3 anemia was found in 6.7% of the patients. Twelve cases (23%) had grade 3 neutropenia and only one patient (2%) had grade 4. There was no thrombocytopenia grade 3 or higher. For late toxicities, five patients had growth hormone deficiency and two patients had hypothyroidism which required medical treatment. Six patients had high frequency hearing loss.

## Discussion

The patient characteristics of our study were similar to other studies including median age of 7 years (IQR 5-10), and predominance of male, cerebellar vermis location and classic histology. [10, 14, 15] The majority of the cases in our study were classified as nonWNT/nonSHH subtype which was in agreement with other studies [4, 11, 16]. The correlation between molecular findings and histological subtypes was also in line with many studies [13, 17, 18]. One case from the SHH subgroup had desmoplastic/nodular histology while all WNT and nonWNT/nonSHH subgroups had classic histology.

The 5-year overall survival rates of MB in our study were 76.3% for the standard-risk group and 71.4% for the high-risk group. These findings were in line with previous studies that reported the 5-year overall survival rates of MB to be 70-80% and 60-70% for standard-risk group and high-risk group, respectively [3-5]. In contrast, a previous study conducted in Thailand in 55 pediatric MB patients had a 5-year OS rate of 84.4% in the standard-risk group and 42.8% in the high-risk group; [10] for this study, the survival rate in the high-risk group was higher than the previous Thai study. This could probably be due to higher gross tumor resection rate, higher rate of combination chemotherapy with radiotherapy, and also higher rate of craniospinal irradiation in our study. In our previous study, [19] MB patients treated between 1989 and 1997 had a 5-year OS rate of 57.9%. In contrast, this study had a higher 5-year OS rate which may be due to the advancement of the surgical procedures, use of modernized radiotherapy techniques and systemic chemotherapy; all of these treatment strategies have improved the treatment outcome.

Post-CSI radiation treatment target volume in MB can be performed in two ways, either involved field or posterior fossa radiation therapy. The ACNS 0331 study reported that the involved-field RT was as effective as posterior fossa RT of 54 Gy following CSI in the standard risk MB group [20]. However, whether this is also true for high-risk patients is still inconclusive. In our study, there were 14 cases and 13 cases in the high-risk group receiving posterior fossa boost and tumor bed boost, respectively. Local relapse was recorded in 3 out of 14 and 3 out of 13 patients in each treatment group. This suggests that tumor bed boost following CSI of 36 Gy could be a treatment option in high-risk MB patients.

With improvement of survival rate in pediatric MB patients, long-term toxicities are of concerns for the survivors. Current clinical trials are focusing on identification of low-risk WNT subgroup which has a potential for treatment de-escalation. Although, our analysis showed no significant difference in survival between the molecular subtypes, our two patients with WNT subtype had very good outcome with 100% 5-year OS rate. This was in agreement with previous studies which reported that the WNT subtype had the best prognosis [21, 22]. Therefore, molecular data has been used to classify good-risk patients in on-going clinical trials which could reduce treatment-related toxicities and improve quality of life while maintaining the excellent outcome for pediatric MB patients. However, clinical research that incorporated molecular subtype are lacking in Southeast Asia due to the limitations of resources for molecular subtyping. Our study is the first to report the clinical outcomes of MB patients using simplified molecular subtyping. The molecular subtypes were determined by immunohistochemistry which showed that it was reliable and easily obtainable in all laboratories [13].

In the multivariate analysis, the prognostic factor significantly associated with treatment outcome was the time interval between surgery and RT. When it was ≥ 8 weeks, the patients would have poorer 5-year OS compared to those who had less than 8 weeks. Typically, most clinicians try to start RT within 4 weeks after surgery in an effort to prevent tumor regrowth. For example, current clinical trials in MB patients stated that adjuvant RT should be initiated within 31 or 36 days after surgery (NCT00392327 and NCT01878617). However, the optimal timing of the interval remains unclear. Our study is in line with recent National Cancer Database (NCDB) analysis that reported postoperative radiotherapy deferral for more than 90 days after surgery was associated with poorer OS [23]. Moreover, previous randomized studies that compared early RT versus up-front postoperative chemotherapy and delayed RT also demonstrated inferior OS when RT was delayed [24, 25]. On the other hand, in a large National Database analysis [7], early RT (defined as initiation ≤ 3 weeks after surgery) had decreased 5-year OS while time to RT > 5 weeks but within 90 days did not have any adverse survival impact. To the best of our knowledge, there is no large prospective study that investigated the outcome of RT timing after surgery in MB pediatric patients. Our result found no detrimental impact on survival after delaying radiation for more than 4 weeks but within 8 weeks. Most of the cases in our study that had time interval ≥ 4 weeks were due to the delayed referral process from other hospitals that had limited resources to receive radiation at our center. Our analysis suggested that the timing of post-op RT should be given as soon as possible after there is adequate wound healing and multidisciplinary evaluation but not over 8 weeks after surgery.

There were some limitations in our study. First, limitation regarding the immunohistochemistry method cannot separate group 3 and group 4 subtypes. Second, not all patients had molecular subgroup results due to the retrospective nature of the study. Last, the number of patients in our study was small because of the disease rarity and single institution. Therefore, prospective studies of more patients are warranted.

## Conclusions

The 5-year OS rate in our center has improved during the past 3 decades from 57.9 % to 74.2% which currently was consistent with other studies. The only statistically significant prognostic factor was the time interval of up to 8 weeks between surgery and RT. Incorporating simplified molecular studies with clinical characteristics might be helpful in better identification of good-risk patients for future clinical trials.

## Data Availability

To protect potentially identifiable information on serious crimes, ethical approval is needed to access data. Data are available from ethics commitee at faculty of medicine, Chulalongkorn university (contact via) for researchers who meet the criteria for access to confidential data.

## Acknowledgments

We would like to thank the English editing service offered through the Research Affairs at the Faculty of Medicine, Chulalongkorn University. We are grateful to Buntipa Netsawang for her contribution

## Supporting information

**S1 Table. Demographic data, molecular and histology subtypes**.

